# THE ECONOMIC BURDEN OF INVASIVE FORMS OF MENINGOCOCCAL INFECTION

**DOI:** 10.1101/2024.06.25.24309375

**Authors:** E.N. Sergienko, I.N. Kozhanova, O.N. Romanova, A.D. Solodov, P.A. Scutova

## Abstract

**Relevance:** Invasive meningococcal infection (IMI) is a significant clinical problem and is associated with a high probability of severe complications and death. Vaccination programs against meningococcus can achieve significant clinical effectiveness and require significant financial costs. This makes it relevant to study the economic burden of meningococcal infection in the conditions of modern healthcare in order to obtain basic data for subsequent research in the field of evaluation of medical technologies.

**Aims:** To assess the economic burden of IMI in children in the conditions of healthcare in the Republic of Belarus.

**Material and methods:** a retrospective study using the “cost of illness” method was conducted for 22 children hospitalized in the city children’s infectious clinical diseases hospital in 2018-2019. All values are presented in belarusian rubles. The average ± standard deviation, minimum – maximum values, median [Q1, Q3], cost shares (%) are calculated for the cost values. The minimum– maximum values and median [Q1, Q3] were calculated for the age of the patients.

**Results:** taking into account the costs of treatment in healthcare organizations of all patients studied, the share of direct costs of hospitalization was 29.0%, the share of direct costs of diagnosis - 7.2%, the share of direct costs of pharmacotherapy - 8.3%. A significant part of the costs accounted for observations after discharge and amounted to 55.4%.

**Conclusion:** a retrospective study of the economic burden of MI in the Republic of Belarus using the “cost of illness” method in patients hospitalized in a public health organization demonstrates a significant economic burden of this disease, which in the long term is primarily due to the presence of complications in the patient, as well as the impact of the patient’s death on the value of total costs.

## Introduction

Invasive meningococcal disease (IMD) is an infectious disease caused by bacteria called Neisseria meningitidis. Currently, twelve different serogroups of N. meningitidis are known, with more than 96% of IMD cases being attributed to serogroups A, B, C, Y, and W [1]. Despite the availability of antibiotic therapy, IMD poses a significant clinical challenge due to its unpredictable course and severe sequelae. The mortality rate in different countries varies from 10% to 40%, while complications of the disease develop in 10%-30% of survivors [2, 3].

The global annual registration of IMD ranges from 1.2 to 1.4 million cases, resulting in 135,000 to 150,000 fatalities. This disease predominantly affects infants and children under 5 years of age, as well as adolescents and young adults [4,5]. In Europe, in recent years, the overall incidence in the general population is 0.7–0.9 cases per 100,000 population (ranging from 0.4 to 2.5–3.2 per 100,000 population), and is almost 20 times higher in infants under 1 year of age (up to 16 cases per 100,000 children) [6,7]. The widespread carriage of this infection, especially among adolescents and young adults, contributes significantly to its significance. According to some data, the carriage level in this population can reach 25–30%. Carriers are known to be the most epidemiologically significant source of infection. As literature suggests, there are approximately 2-3 thousand carriers of N. meningitidis per 1 patient [8].

Prior to 1913, the mortality rate for IMD was as high as 70-90%. Today, the average mortality rate is 10-20%, being primarily attributed to fulminant forms of IMD. This places meningococcal infection in the first place in terms of mortality among vaccine-preventable infections [9]. Up to 75% of all fatal cases occur in children in the first 5 years of life, with 40% of these cases developing in the first year of life. Mortality depends not only on the age of the patient, but also on the serogroup of the meningococcus that caused IMD. Serogroup W is associated with the highest number of deaths (12.8%), followed by serogroup C (12.0%), serogroup Y (10.8%), and serogroup B (6.9%). Also, the clinical form of the disease plays a significant role, with the highest mortality rates observed in cases of meningococcal sepsis (meningococcemia), especially those with septic shock [7,9].

Up to 20–30% of IMD survivors face complications leading to long-term problems [10]. Complications can include both early neurological problems (seizures, subdural effusion or empyema, hydrocephalus, increased intracranial pressure, focal neurological deficits, cerebral venous sinus thrombosis, cerebral infarction) that can be associated with shock and tissue hypoperfusion (skin necrosis, gangrene of a part of or the entire limb (several limbs)), and long-term neuropsychological problems (deafness, epilepsy, learning difficulties, motor and mental developmental disorders) that can be associated with tissue hypoperfusion (skin scars (skin grafts may be required), growth plate injury (multiple surgical procedures may be required before bone growth is complete), arthritis with or without joint damage, etc.). In addition, it was found that in children under 1 year of age with IMD, the adjusted odds ratio of death was 4.6 times higher compared to the group without the disease. Surviving children experience a significant rise in general disability rates 3-5 years post-illness; patients who had IMD in childhood exhibit markedly reduced average indicators of physical health [11]. In fact, the impact of IMD extends beyond the patient. Often the burden of IMD on the patient’s family, society at large and other aspects (legal matters, social problems, psychological stress, etc.) are neglected, since it is very difficult to evaluate it in monetary terms.

Between 1998 and 2020, the Republic of Belarus witnessed a notable decline in the incidence of IMD (a drop from 3.7 to 0.4 cases per 100,000 population), exceeding the epidemic threshold (more than 2 cases per 100,000 population) before 2008 [12]. However, in 2021 and 2022, there has been a resurgence in the incidence rate, reaching 0.7 and 1.3 cases per 100,000 population, respectively. Similar trends have been observed globally in the post-COVID-19 period. Throughout the years 1998–2020, the population mortality rate ranged from 3.8% in 2008 to 15.7% in 2020, with an average IMD mortality rate of 11.9% during this period [12]. IMD cases are reported in all regions of the Republic of Belarus throughout the year, with the epidemiological process intensifying in the spring. The serogroup distribution of meningococci causing IMD in different regions of the country exhibits distinct characteristics. While meningococcal serogroup B dominates nationwide, an analysis spanning the past decade reveals a 50% increase in the proportion of serogroups A, C, W, and Y [12].

There is a notable absence of available data in the Republic of Belarus concerning long-term outcomes, healthcare resource utilization, and the long-term economic burden of IMD, which makes this study relevant.

The study purpose is to assess the economic burden of IMD in children in the healthcare settings of the Republic of Belarus.

## Materials and methods

The economic evaluation was carried out in a prospective research study to establish the characteristics of meningococcal sepsis in children, taking into account the course of the disease. The clinical part of the study followed a prospective design (with tracking of long-term sequelae), while the economic part adopted a retrospective approach utilizing the “cost-of-illness” methodology [13]. The economic burden study used data from 22 children with IMD consecutively admitted to the municipal children’s infectious diseases clinical hospital in 2018–2019. Demographic information on patients (sex, age), use of health care resources in the management of each patient, information on complications and long-term sequelae of the disease (four-year follow-up) were collected. The “cost-of-illness” analysis encompassed direct medical costs of hospitalization (hospitalization itself, diagnosis, drug therapy); direct medical costs of infection-related sequelae (amputation, cochlear implantation, post-discharge outpatient follow-up); indirect costs (payments of temporary disability benefits to a parent of children aged 3 to 14, gross domestic product (GDP) lost due to temporary disability of a parent of children aged 3 to 14 and lost years of productive life due to the death of the patient). All financial calculations were made in 2022–2023 using current tariffs. All figures are presented in Belarusian rubles. For cost values, the mean ± standard deviation, minimum – maximum values, median [Q1, Q3], cost shares (%) were calculated. The minimum and maximum values and median [Q1, Q3] were calculated for the age of the patients.

### Study results

For this study, data were gathered from 22 patients, with a median age of 1.67 years (ranging from 0.83 to 3 years). The youngest child was 1 month old; the oldest was 17 years old. The majority (86.4%) were children under 5 years of age, with girls accounting for 36.4% and boys for 63.6%. Of the 22 cases, 19 (86.4%) patients had a mixed form of IMD (meningococcemia, purulent meningitis) and 3 patients (13.6%) had purulent meningitis of meningococcal etiology. Septic shock developed in 5 patients (22.7%) and one patient (4.54%) had Waterhouse-Friderichsen syndrome. The average length of stay in the hospital was 16.59±6.23 days (range: 7 to 29 days). Of these, length of stay in the intensive care unit was 5.77±3.62 days (range: 2 to 13 days). Length of stay in an infectious diseases department ward averaged 11.8±5.98 days (range: 2 to 23 days). Discharge outcomes included recovery in 18.2% of children, improvement in 68.2%, transfer to another institution in 9.1%. One child died. Two patients underwent cochlear implantation due to sensorineural hearing loss; one patient underwent lower extremity amputation. Table 1 provides details on the direct medical costs for pediatric patients with IMD.

**Table 1.** Characteristics of direct medical costs in children’s patients with IMI, BYN/US dollars.

| Cost type | Median | Interquartile range |  | Minimum value | Maximum value |
| --- | --- | --- | --- | --- | --- |
| **Direct medical costs** | 8 935,00/<br>2 792,2 | 5 662,56/<br>1 769,6 | 12 903,31/<br>4 032,3 | 2 899,4/<br>906,1 | 93 744,7/<br>29 295,2 |
| **of these, the direct costs of hospitalization** | 5 002,98/<br>1 563,4 | 3 860,28/<br>1 206,3 | 6 485,83/<br>2 026,8 | 2 313,4/<br>722,9 | 9 339,6/<br>2 918,6 |
| **of these, direct diagnostic costs** | 1 107,64/<br>346,1 | 718,40/<br>224,5 | 1 621,68/<br>506,8 | 394,0/<br>123,1 | 3 029,3/<br>946,7 |
| **of these, the direct costs of pharmacotherapy** | 534,81/<br>167,1 | 195,12/<br>60,9 | 1 884,98/<br>589,1 | 49,9/<br>15,6 | 12 283,2/<br>3 838,5 |
| **follow-up after discharge** | 882,53/<br>275,8 | 882,53/<br>275,8 | 1 000,13/<br>312,5 | 882,53/<br>275,8 | 84 900,0/<br>26 531,3 |
| The share of each type of cost in the total direct medical costs per 22 patients |  |  |  |  |  |
| **Share of direct hospitalization costs, %** |  |  |  | 29,0 |  |
| **Share of direct diagnostic costs, %** |  |  |  | 7,2 |  |
| **Share of direct pharmacotherapy costs, %** |  |  |  | 8,3 |  |
| **Share of follow-up after discharge** |  |  |  | 55,4 |  |

Direct medical costs for the management of patients with childhood IMD will amount to a median of 8935 BYN (5662.56–12903.31 BYN), including direct costs of hospitalization – 5002.98 BYN (3860.28–6485.83 BYN); direct costs of diagnosis – 1107.64 BYN (718.40–1621.68 BYN); direct costs of drug therapy 534.81 BYN (195.12– 1884.98 BYN) (Figure 1).

**Figure 1.**
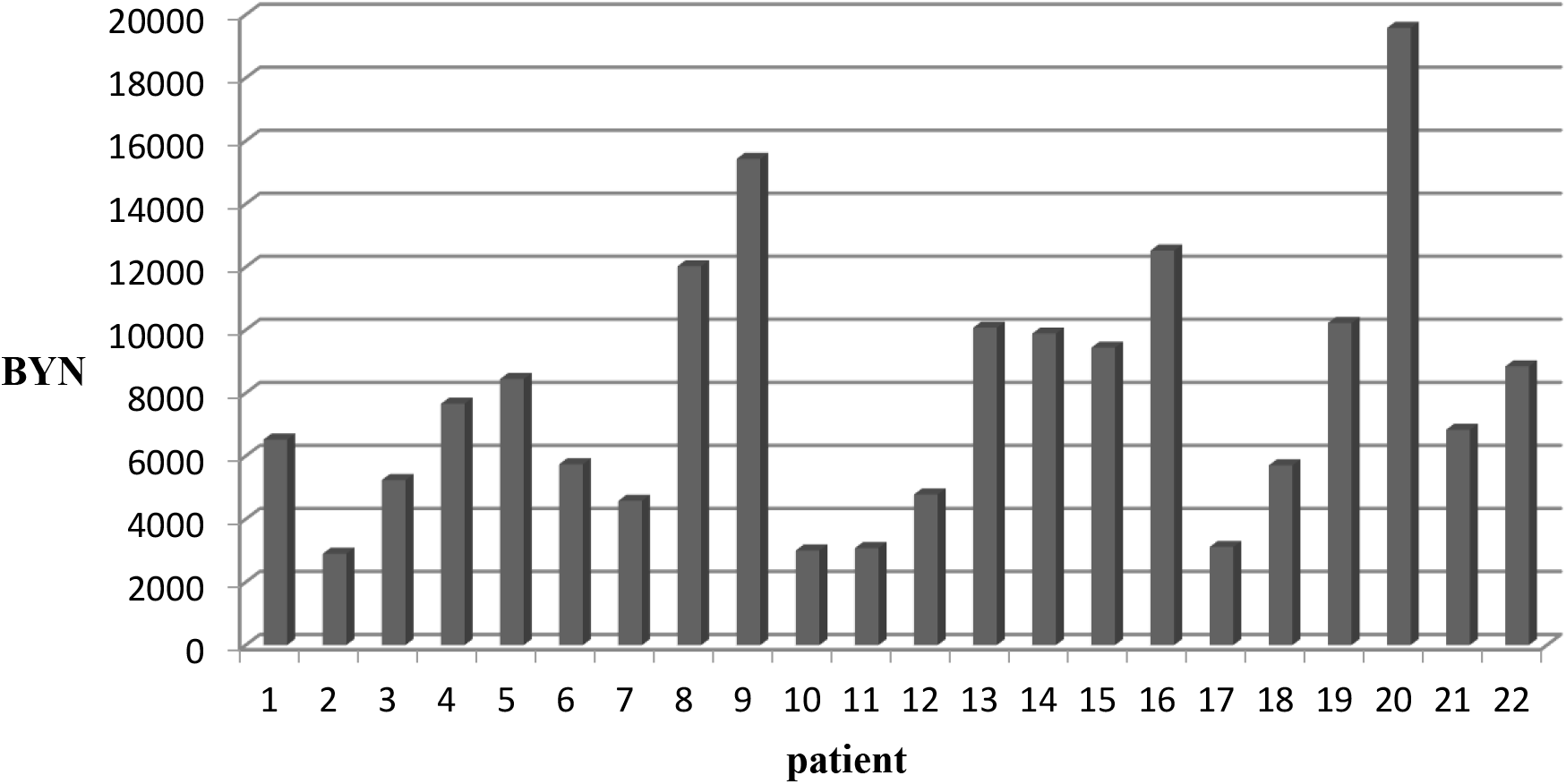
Direct medical costs for the acute phase of the disease (hospitalization), BYN.

Patients post IMD require post-discharge follow-up, which includes examinations by specialists (pediatrician, infectious disease specialist, neurologist, ophthalmologist, audiologist) and instrumental investigations (EEG, transcranial Doppler ultrasound, MRI, neurosonography). Also, several cases of severe complications of the infection were reported in patients included in the study: amputation of the lower extremity, two cases of sensorineural hearing loss requiring cochlear implantation. Accordingly, direct medical costs of post-discharge patient follow-up amounted to a median of 882.53 BYN (882.53 – 1000.13 BYN), with minimum and maximum costs ranging from 882.5 BYN to 84900.0 BYN per patient. In general, taking into account the costs of treatment in healthcare organizations for all studied patients, the share of direct costs of hospitalization was 29.0%, the share of direct costs of diagnosis was 7.2%, and the share of direct costs of drug therapy was 8.3%. A significant portion of the costs accounted for post-discharge follow-up and amounted to 55.4% (Figure 2).

**Figure 2.**
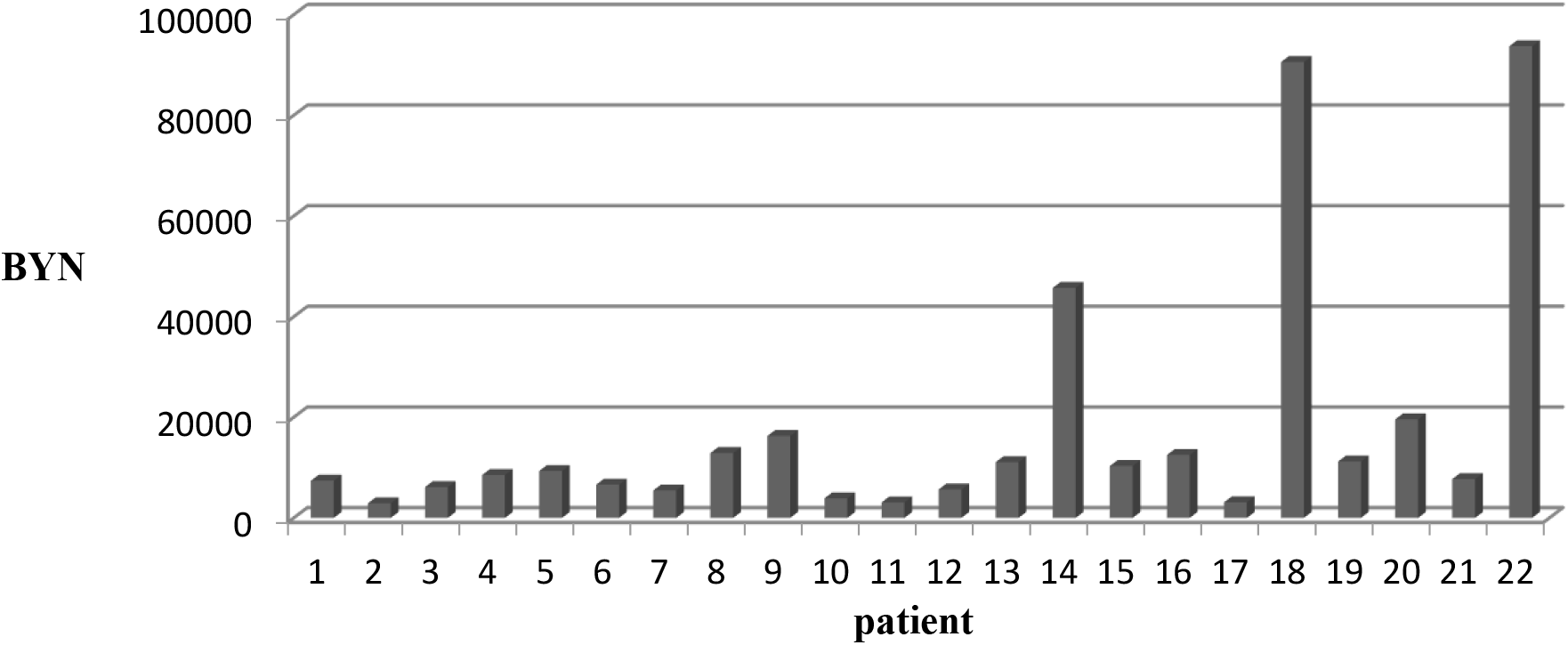
Direct medical costs in providing care to patients with IMI, taking into account follow-up after discharge and complications that have developed, BYN.

The indirect per-patient costs (not directly related to the health care services) included: temporary disability payments – a median of 933 BYN (882.5–1152.96 BYN); GDP losses during the temporary disability period – 2191 BYN (1773.4–2503.6 BYN). Death of a patient, particularly in terms of the working age perspective, contributed significantly to indirect costs, amounting to 930,579 BYN (Figure 3).

**Figure 3.**
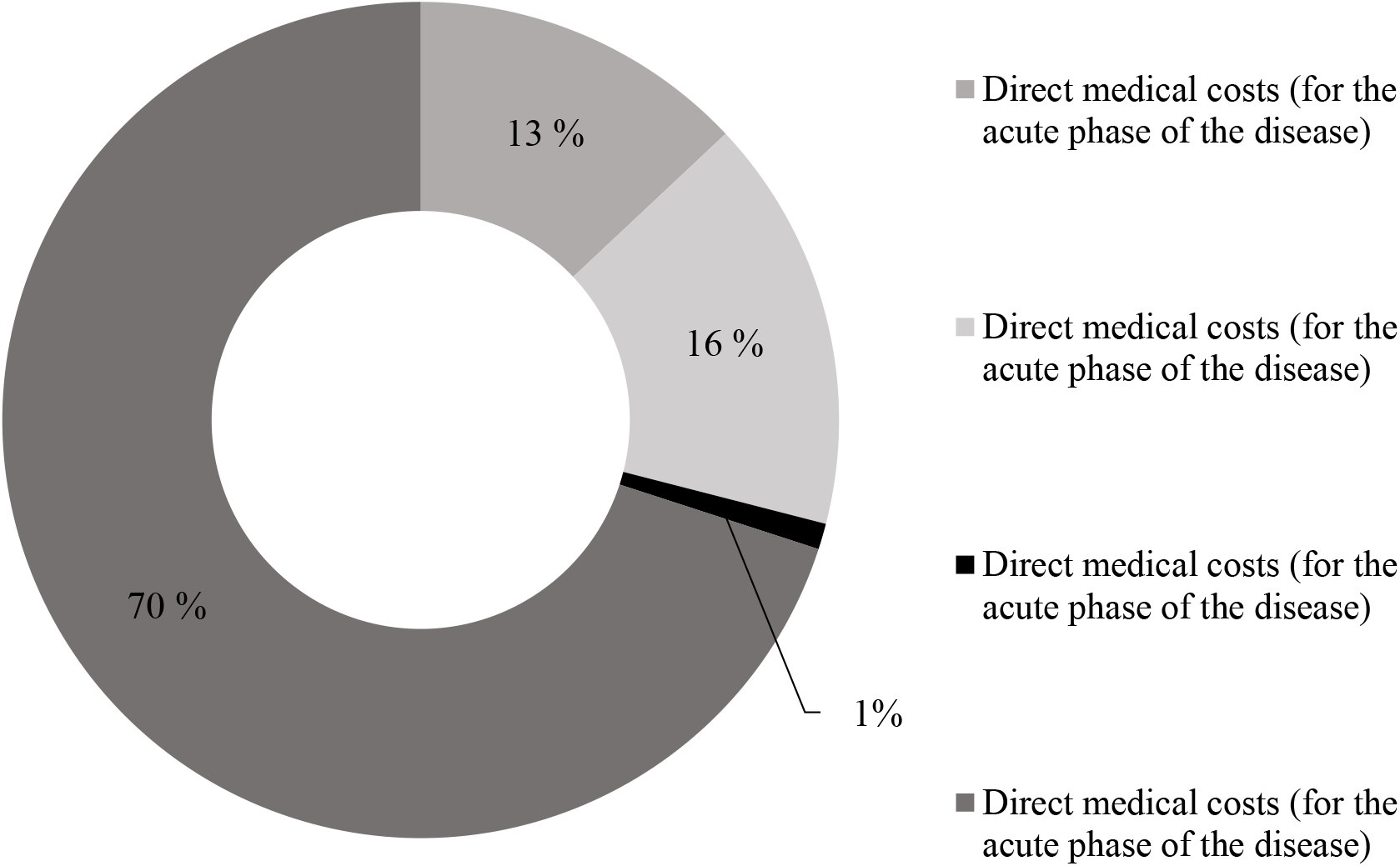
Cost structure for the study cohort of patients (22 people) with IMI.

Taking into account the total costs for all patients included in the study, direct medical costs constituted 29.37%, while indirect costs amounted to 70.63% of the total. In absolute values, direct medical costs per patient are 17,902 BYN, total costs per patient are 60,948 BYN.

The study involved 22 patients, with three experiencing IMD sequelae such as limb amputation and cochlear implantation in two cases. One patient died, which was also assessed in the analysis as the infection-related sequela. This group of patients (with infection-related sequelae) contributes a significant disproportion to the economic burden of the disease (Figure 4).

**Figure 4.**
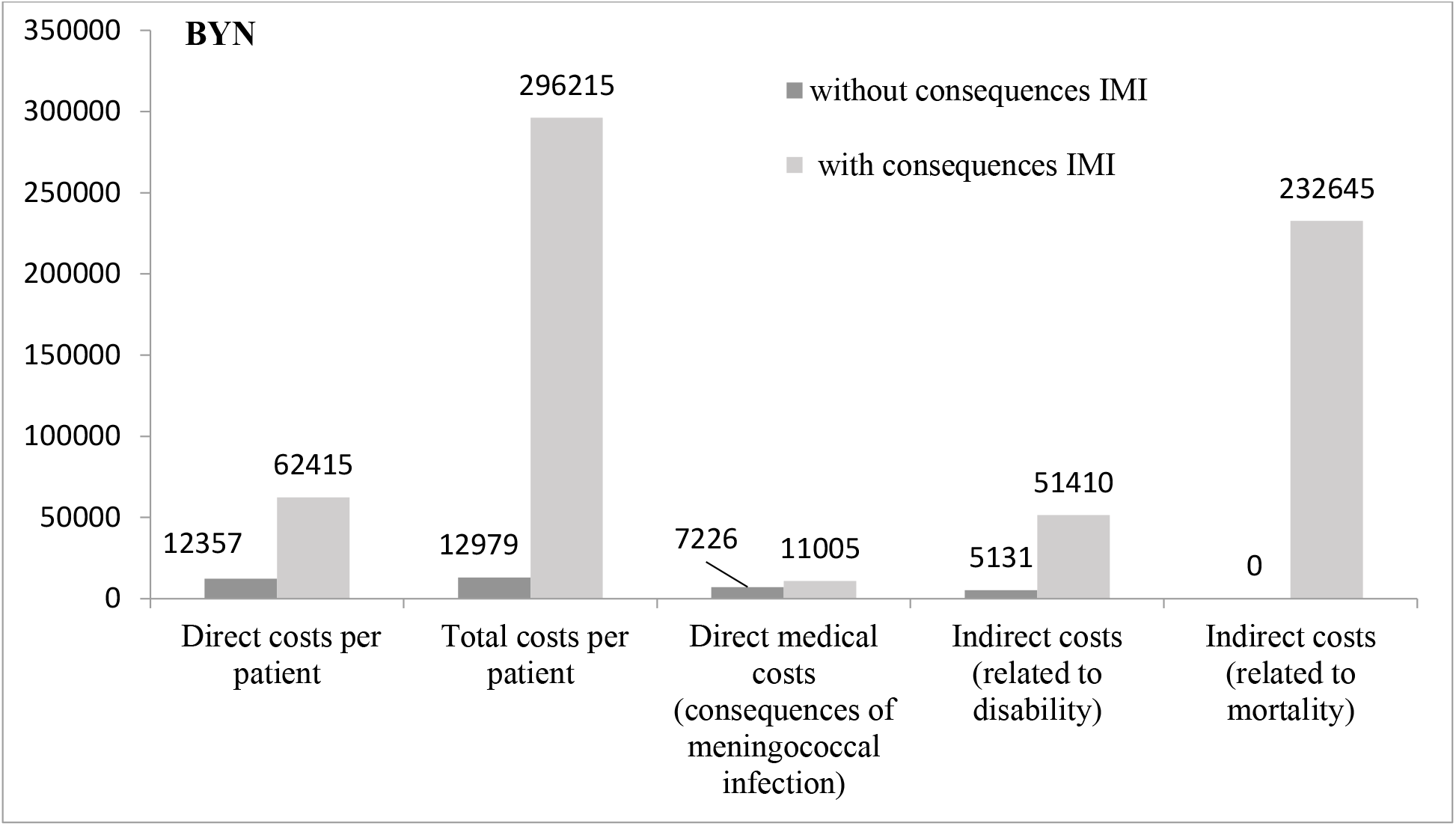
Cost characteristics in patients with and without consequences of IMI, BYN.

Thus, in patients with sequelae of IMD, direct medical costs were more than five times higher than in patients without sequelae, and total costs were more than 22 times higher for all individual types of costs: direct medical costs for the acute phase of the disease were 1.5 times higher, direct medical costs for sequelae of IMD were 10 times higher. Premature death makes a significant contribution to the economic burden of IMD and its sequelae.

## Discussion

The study shows that although IMI is not a widespread disease, it imposes a substantial economic burden on both the healthcare system and society as a whole. In absolute terms, direct medical costs per patient are 17,902 BYN, total costs per patient are 60,948 BYN. Taking into account the total costs for all patients included in the study, direct medical costs constituted 29.37%, while indirect costs amounted to 70.63% of the total. Patients with complications of IMD, as well as those succumbing to the infection, make a significant and disproportionate contribution to the costs, increasing them tenfold compared to patients without complications. This is noteworthy, as post-infectious complications are a distinctive feature of meningococcal infection. Thus, according to literature, 23.5% to 58% develop sequelae of IMD [14-16].

In the study by Huang (2020) [17], the probability of developing complications was 13% at hospital discharge and 23.5% in the subsequent follow-up period. In a systematic review, Strifler (2016) [18] found that the most common sequelae of IMD were hearing loss, cognitive impairment, and psychological problems. The most common complications were Waterhouse-Friderichsen syndrome, accompanied by adrenal hemorrhage (11.6%), and chronic renal failure (7.5%). A study by Davis (2011) [14] analyzed the incidence of IMD complications over one year after the initial diagnosis. Chronic renal failure was observed in 9% of IMD cases, but no cases of adrenal hemorrhage were observed. Among the patients included in our study, one patient underwent amputation (4.54%), two patients required cochlear implantation (9.09%), and one patient died (4.54%). Waterhouse-Friderichsen syndrome was reported in one patient (4.54%).

The economic burden of meningococcal infection has been analyzed in numerous foreign studies. Direct comparisons with our study are challenging due to the differences in the design, health care and welfare financing systems, and the basic economic indicators used to estimate the long-term sequelae of the disease. At the same time, the results of these studies confirm the trends we noted: a significant economic burden of an acute condition, disproportionality of costs associated with the presence/absence of disease-related sequelae, the predominance of indirect medical costs and costs related to sequelae.

The study by Weil-Olivier (2021) [19] found that the average cost of hospitalization per person was €11,256 and increased for patients with complications. Annual post-IMD costs were €4254 in cases without sequelae, €10,799 euros in cases with one sequela and €20,096 euros in cases with several sequelae. Additional costs were primarily associated with the management of disease-related sequelae. Among sequelae, the authors noted amputations, skin scarring and mental retardation, which resulted in per-patient costs exceeding €20,000 in the first year and over €10,000 in the subsequent years.

The German study by Huang (2020) [17] showed that the average cost of hospitalization associated with IMD was €9,620 ± €22,197. IMD resulted in severe complications and sequelae and was associated with higher utilization of health care resources.

Health care costs have been estimated at C$45,768–52,631 per case of IMD in Canada, US$23,294 per hospitalization in the United States, and US$18,920 per case in Denmark [20].

The Mexican study of IMD-related hospitalizations by Chacon-Cruz (2022) [20] showed that the average cost per case was US$20,676, of which direct healthcare costs were US$20,195 and societal costs were US$481.

The costs of acute episodes of IMD were estimated in a study by Christensen (2013) [21] and amounted to 5045 and 4798 euros per case of IMD in patients aged <16 years and >16 years, respectively. A similar result was found in an Australian study, where the average costs of hospitalization for acute IMD were significantly higher compared with the costs of the “average” infectious patient in public hospitals (US$12,312 versus US$4,918, respectively) [15].

A French study by Bénard (2016) [22] assessed the lifetime costs of severe IMD per case from a societal perspective. Lifetime costs ranged from €768,875 to €2,267,251 depending on expected complications and sequelae.

It is important to mention certain methodological limitations of the study. The data were derived from a single healthcare institution. Assessment of costs did not account for societal costs (costs of care, payments for probable disability, financial losses of the family due to psychological problems associated with the child’s illness) with the exception of payments for temporary disability and GDP losses.

## Conclusion

The retrospective study of the economic burden of IMD in the Republic of Belarus using the “cost-of-illness” method in patients hospitalized in a public health care institution demonstrates the significant economic burden of this disease, which over the long term is primarily due to complications in patients, as well as the impact of a patient death on total costs. The data obtained can be used to conduct national medical technology assessment studies, particularly in clinical and economic modeling. In particular, such studies are essential for assessing the feasibility and selection of vaccination programs and other preventive measures, as well as for justifying optimal strategies for managing patients with an acute illness [23,24].

Hence, IMD-related costs vary depending on a number of factors, including clinical manifestations of the disease and the presence/absence of long-term sequelae in patients. The economic burden of IMD varies across countries, contingent on the availability of health care resources, financing principles, and characteristics of the national economy. Understanding the national burden of IMD and its broad economic impact (beyond direct medical costs directly for the public health care) has enabled informed decision-making regarding resource allocation priorities.

**No conflict of interest to declare**.

## Data Availability

All data produced in the present study are available upon reasonable request to the authors

